# Octopi 2.0: Open and Scalable Microscopy Platform for AI-powered Diagnostic Applications

**DOI:** 10.1101/2025.03.21.25324364

**Authors:** Hongquan Li, Heguang Lin, Pranav Shrestha, Rinni Bhansali, You Yan, Jaspreet Pannu, Kevin Marx, Wei Ouyang, Lucas Fuentes Valenzuela, Ethan Li, Anesta Kothari, Jerome Nowak, Hazel Soto-Montoya, Adil Jussupov, Maxime Voisin, Kajal Maran, Oswald Byaruhanga, Joaniter Nankabirwa, Bryan Greenhouse, Prasanna Jagannathan, Manu Prakash

**Author notes:** Work done during an internship at Cephla Inc (Heguang Lin).

## Abstract

Access to quantitative, robust, and affordable diagnostic tools is essential to address the global burden of infectious diseases. While manual microscopy remains a cornerstone of diagnostic workflows due to its broad adaptability, it is labor-intensive and prone to human error. Recent advances in artificial intelligence (AI) and robotics offer opportunities to automate and enhance microscopy, enabling high-throughput, multi-disease diagnostics with minimal reliance on complex supply chains. However, current automated microscopy platforms are often costly and inflexible — barriers that are especially limiting in low-resource settings. Here we present Octopi 2.0, an open, highly configurable, general-purpose automated microscopy platform for a broad range of diagnostic applications, including sickle cell anemia and antibiotic resistance that we have reported recently. Applying Octopi to imaging malaria parasites with 4’,6-diamidino-2-phenylindole (DAPI) staining, we discovered a spectral shift in fluorescence emission that allows rapid screening of blood smears at low magnification with throughput on the order of 1 million blood cells per minute. We further developed image processing and deep learning-based segmentation and classification pipelines to enable real-time processing for malaria diagnosis. For real-world performance validation, we collected a data set of 213 clinical samples from Uganda and the United States with a total of 905 million red blood cells and around 1.4 million malaria parasites. Using a ResNet-18 model and only one round of retraining, the model is able to achieve on average less than 5 false positive parasites/µL and a per-parasite level false negative rate of less than 8% in our test dataset. This per-cell performance implies a limit of detection (LoD) around 12 parasites/µL, and we measured patient-level performance of *>*97% specificity and sensitivity in our independent test data set of clinical samples from 73 patients/donors. As more data is collected in larger validation studies, we expect the robustness and performance of the model to continue to improve according to what we observe in our proof-of-concept experiments carried out in this study. With significant cost reduction in hardware compared to current automated microscopes and an open and versatile approach for tackling multiple diseases with standard glass slide-based sample preparation, we envision Octopi 2.0 to help enable the “app store” for equitable data-driven, AI-powered diagnostics of many diseases and conditions.

## Introduction

With its role in direct visual identification of pathogens and pathological conditions, microscopy remains a WHO gold standard for numerous diseases [1]. However, despite technological advancements in related fields, the practice of microscopy has remained largely unchanged over the last half century, especially in low- and middle-income countries [2]. Moreover, health systems have not been able to implement microscopy at a scale that meets the needs of the communities they serve [3].

Microscopic detection of malaria parasites is one of the leading applications of microscopy-based diagnosis, with huge unmet needs. Nearly half of the world’s population is at risk of malaria [4, 2, 5]. In the year of 2023, there were 263 million malaria cases, up from 249 million in 2022, and nearly 597,000 deaths, the majority of which is attributable to *Plasmodium falciparum*, a strain of malaria widely spread across the world [3, 4]. The two most widely used diagnostic tests are antigen-based rapid diagnostic test (RDT) and microscopic examination of blood smears. In the same year (2023), 348 million RDTs were used and more than 217 million patients were tested by microscopy, whereas the estimated need for testing was well over 1 billion [4, 2, 6].

RDTs have significantly expanded test availability but have the following limitations [7]: (1) the most widely used RDTs are not quantitative and cannot be used to monitor treatment response. (2) RDTs can remain positive for several weeks after parasite clearance. (3) Currently the most sensitive RDTs are based on the HRP2 antigen. However, HRP2 and HRP3 gene deletions make these RDTs unreliable in regions where mutated strains are spreading [3, 7].

While microscopy remains the gold standard for routine diagnostics, “the quality of microscopy-based diagnosis is frequently inadequate” [8]. According to a recent study [9], the sensitivity of manual or expert microscopy is only 30% at parasitemia of 100 parasites per *µ*L and 60% at 1000 parasites per *µ*L, with large variations across clinics and individuals. These numbers should be compared to the estimated detection limit of expert microscopy, which is below 50 parasites per *µ*L. Manual microscopy is also labor-intensive, as an expert microscopist needs to spend around 6 minutes to examine a single slide that is negative or has low parasitemia [10]. Furthermore, manual microscopy requires extensive training and quality assurance programs, much of which is poorly or not implemented in resource-limited settings [11]. Commercial slide scanning and flow cytometry systems show promise [12, 13, 14, 15, 16, 17] but have not become mainstream, where the high cost of capital equipment and in some cases, use of proprietary consumables are likely hindering factors. With persisting high burden of malaria [18] and the rise of mutated strains [19, 20, 21, 22, 23], affordable, high-throughput and quantitative diagnostic tests are urgently needed.

In the past decade, low-cost and portable microscopes have made tremendous strides [24], both towards improving access and implementing application-specific capabilities such as disease diagnosis using field-portable microscopes [25, 26, 27, 28, 29, 30, 31, 32, 33, 34, 35]. These platforms and techniques have demonstrated a wide range of applications [36, 37, 38], including microscopic detection of malaria-infected red blood cells [32, 39]. However, accurate (matching or surpassing the performance of expert microscopy) and high-throughput diagnosis of malaria has remained out of reach, either due to the long scanning time needed or insufficient resolution for highly specific detection. In the meantime, advances in machine learning have fueled the development of automated parasite detection pipelines for high magnification images of Giemsa-stained blood smears [40, 41, 42]. Yet the level of per-cell specificity achieved in many of the reports is still not sufficient for diagnosis at very low parasitemia - even per-cell specificity of 99.9% would still mean around 5000 healthy RBCs per *µ*L of blood will be falsely identified as infected, hindering their use in early detection and treatment monitoring. Recent efforts leveraging 405 nm transillumination have shown promising results [43], but reliance on customized cartridges increases cost and can limit reach.

Here we present Octopi 2.0, a cost-effective, portable, AI-powered, modular, and automated imaging platform designed for easier, accurate, and highly scalable implementation of microscopic disease diagnosis and treatment monitoring in resource-constrained settings as well as for clinical research and epidemiological studies. As a motorized microscope for standard glass slides, Octopi has already shown promising results in sickle cell diagnostics [44]. Applying Octopi to malaria diagnosis, here we discovered a spectral shift on the order of 10 nm in *P. falciparum* malaria parasites stained using 4’,6-diamidino-2-phenylindole (DAPI), which we take advantage of in rapidly screening red blood cells for infection at throughput on the order of 1 million cells per minute using a 20x/0.5 objective - 100 times faster compared to traditional manual microscopy [45]. Octopi presents a general purpose platform for delivering AI-based diagnostics for multiple pathogens including malaria, tuberculosis (TB) and conditions like sickle cell disease - creating a potential for highly programmable and versatile app store for diagnostics.

## Results

### Open and scalable automated microscopes

Building on our previous work [46, 47], we designed and implemented hardware modules, software, and a framework to build open and scalable automated microscopes (Octopi 2.0) that can be tailored to different diagnostic applications (Fig. 1). The current implementation consists of an LED matrix-based transillumination module that supports differential phase contrast (DPC), a motorized cross-roller bearing XY stage with kinematic mounted glass slide holder, an imaging optical train (infinity-corrected objective lens, tube lens, CMOS camera and depending on configurations, additional optics like dichroics, emission filters and epi-illuminators), a motorized cross-roller bearing z-stage that moves the objective lens or the entire optical train, and a controller (including custom LED driver and stepper motor driver daughter boards) for controlling illumination, motion and hardware trigger of the camera. While cost of standard objective lens can range from tens to thousand(s) of USD, material cost for the rest of the microscope in its basic bright-field only configuration (prototypes) is within $2,000, making the platform more scalable compared to commercial automated microscopes and slide scanners out there. To date, we have applied Octopi platform for detecting malaria parasites (Fig. 1c) in clinical samples, imaging bacterial vaginosis (Fig. 1d) and pap smears (Fig. 1e), sickle cell disease screening (Fig. 1f) [44], and Raman spectroscopy-based detection of bacteria resistance to *β*-lactam antibiotics [48] as well as probing of tuberculosis drug resistance [49]. With its modular and configurable design and high-throughput imaging capabilities, we are working on generating large open-access datasets for training ML models and towards deployment of AI-powered diagnostic solutions (Fig. 1h).

**Figure 1.**
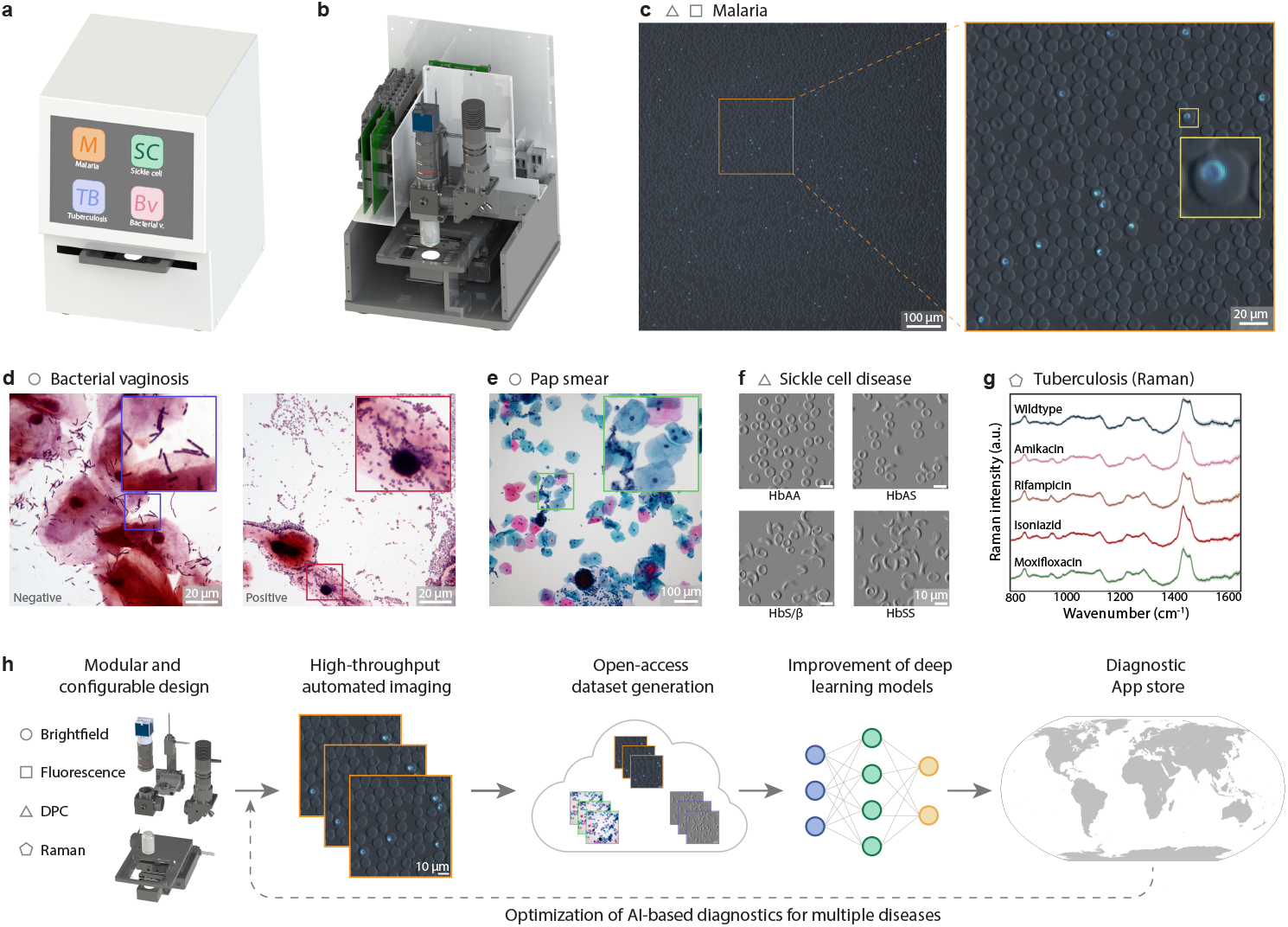
Octopi 2.0 as a scalable platform and an App store model for AI-powered diagnostics. **a-b**, CAD rendering of the Octopi microscope **c**, Example field of view of a DAPI-stained thin blood smear imaged on Octopi with a 20x/0.5 objective (overlay of differential phase contrast and fluorescence). **d**, Example field of view of bacterial vaginosis negative (left) and positive (right) samples imaged on Octopi with a 100x/1.25 oil immersion objective. **e**, Example field of view of pap smear imaged on Octopi with a 20x/0.5 objective. **f**, Images of red blood cells (RBCs) for normal (HbAA), sickle cell trait (HbAS), and sickle cell disease (HbS/*β* and HbSS) participants obtained using Octopi with a 20x/0.5 objective for RBC morphology based classification. **g**, Raman spectra of Mycobacterium tuberculosis (TB) resistant to 4 common anti-TB drugs (Amikacin, Rifampicin, Isoniazid, Moxifloxacin) and pan-susceptible wildtype strain. **h**, Octopi as an open and scalable platform can accelerate development and deployment of AI-powered microscopy-based diagnostics.

### Use of DAPI for staining malaria parasites

Fluorescence microscopy has been used for sensitive detection of malaria parasites [50, 51, 52, 53]. However, the prospect of detection in fixed blood smears at low magnification for increased throughput is hindered by the presence of brightly stained platelets, which are abundant and can appear similar in size and brightness as malaria parasites. For example, a recent work that uses DAPI staining and low magnification for parasite detection still yields around 500 parasites/*µ*L through visual analysis and 2500 parasites/*µ*L through deep learning algorithms for negative samples [32]. Yet *P. falciparum* malaria parasites, which have a 48-hour asexual life cycle, contain not only DNA but also a large amount of RNA. This provides an opportunity for differential detection via a molecular probe. Acridine Orange has been used for differential staining of DNA and RNA and in detecting malaria parasites, however presence of staining artifacts has been identified as a problem [54, 52]. Previously, it has been shown that the emitted fluorescence undergoes a red-shift in DAPI-RNA complexes compared to DAPI-DNA complexes [55]. This property has been used in enumerating reticulocytes in rodent malaria models [56]. We first observed this red shift in DAPI-stained malaria parasites that resulted in parasites and platelets appearing in different colors (green vs blue) on Octopi with 405 nm epifluorescence excitation, a 435 nm long-pass emission filter, and an IMX219 Raspberry Pi camera (color CMOS) using the reversed lens configuration [46]. We subsequently imaged the parasites at higher magnification and with two excitation/emission combinations to confirm the red-shifted emission is primarily from the RNA-rich cytoplasm of the parasites (Fig. 2a and supplementary videos 1 and 2). The manifestation of spectral shift in color is shown through simulation in Fig. 2b.

**Figure 2.**
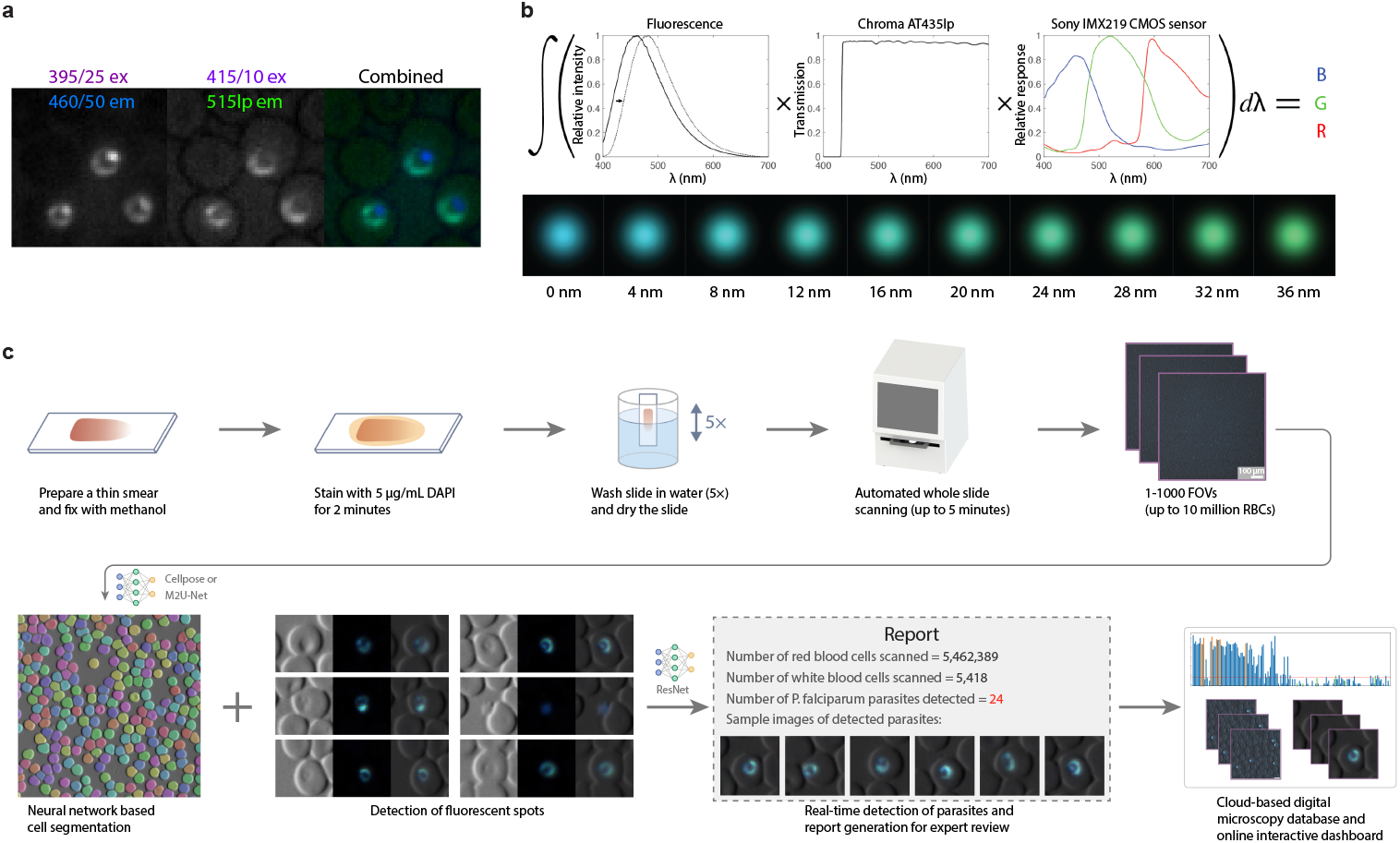
Detection of DAPI-stained malaria parasites. **a**, Images of ring stage Plasmodium falciparum obtained with (1) 395/25 nm excitation and 460/50 nm emission and (2) 415/10 nm excitation and 515 nm long pass emission show DAPI differentially stain nucleus and cytoplasm. **b**, When imaged using a long pass filter and a RGB sensor, the red shift in DAPI emission leads to a difference in color (shown are simulated spots with 0-36 nm red shift). **c**, Procedure for applying Octopi to malaria diagnosis. The processing pipeline implemented on board includes neural network based segmentation of red blood cells, detection of fluorescent spots and image classification using a ResNet18 model. Images of detected parasites will be presented for review and the results can be automatically uploaded for quality control (QC) and real-time surveillance.

Besides differential staining of DNA and RNA, DAPI has several attractive properties, including 20-fold fluorescence enhancement upon binding to the AT-region of dsDNA, low reagent cost (staining a blood smear can cost less than $0.05), short staining time (2 min as compared to 8-10 min for Giemsa at 10% stain working solution and 45-60 min for Giemsa at 3% stain working solution), and good stability - we got similar staining results with DAPI solutions (at staining concentration of 5 *µ*g/mL) left in the dark at room temperature (20 degree C) for a year (Fig. S1).

### High-throughput detection of malaria parasites

Fig. 2c shows the workflow of using Octopi 2.0 with a 20x/0.5 Plan Fluor objective for detecting malaria parasites in DAPI stained blood smears (supplementary video 3). After loading a DAPI stained methanol-fixed thin smear, the microscope will automatically scan a preset region or until a certain number of parasites have been identified. At each location, the microscope takes a fluorescence image and a pair of brightfield images with illuminations from different halves of the LED matrix that are used to generate a differential phase contrast (DPC) image. Red blood cells in the DPC image are segmented and cells that contain fluorescent spots are cropped and passed to a neural network for classification (supplementary video 4). Using a training set consisting of segmentation masks produced by Cellpose [57, 58, 59], we trained an RBC segmentation model with a modified M2U-Net architecture [60]. Our resulting model achieved processing-limited throughput of around 1.5 M RBC/min on laptops with i7 CPU and RTX4070 mobile GPU (around 9,000 RBCs per FOV and 0.34 s per FOV - 20 times faster compared to Cellpose). For slides prepared by Inkwell [61], 13-16M RBCs can be imaged on a single slide. Combining Inkwell smear prep and Octopi slide scanning leads to orders of magnitude increase of data that can be analyzed in reasonable amount of time (tens of seconds to minutes) compared to traditional methods (supplementary video 5). Post scanning, the number of detected parasites, number of red blood cells imaged and parasitemia are reported with images of infected red blood cells displayed for review. These results can be automatically uploaded to cloud-based storage for reporting, quality control (QC) and expansion of a centralized dataset for refining the model to improve its diagnostic performance.

### Data from patient samples for training machine learning models

To evaluate the performance of using Octopi 2.0 to detect malaria parasites, we prepared samples from 96 individuals with 46 malaria positive samples from studies conducted at Infectious Disease Research Collaboration (IDRC) in Uganda and 50 malaria negative samples from IDRC and Stanford Blood Center. A total of 213 thin smears (72 malaria positive and 141 malaria negative) were stained, scanned and processed. Fig. 3a shows the predicted parasitemia in a subset of these slides using a preliminary convolutional neural network binary classifier. The top 10 images of red blood cells with fluorescent inclusions ranked by the classifier from 6 malaria positive slides and 6 malaria negative slides are shown in Fig. 3b. Sampling images from uniform manifold approximation and projection (UMAP) [62] of 3 malaria positive samples, Fig. 3c shows various appearances of malaria parasites within and across samples (UMAP exploration of 10 patient samples in supplementary video 6).

**Figure 3.**
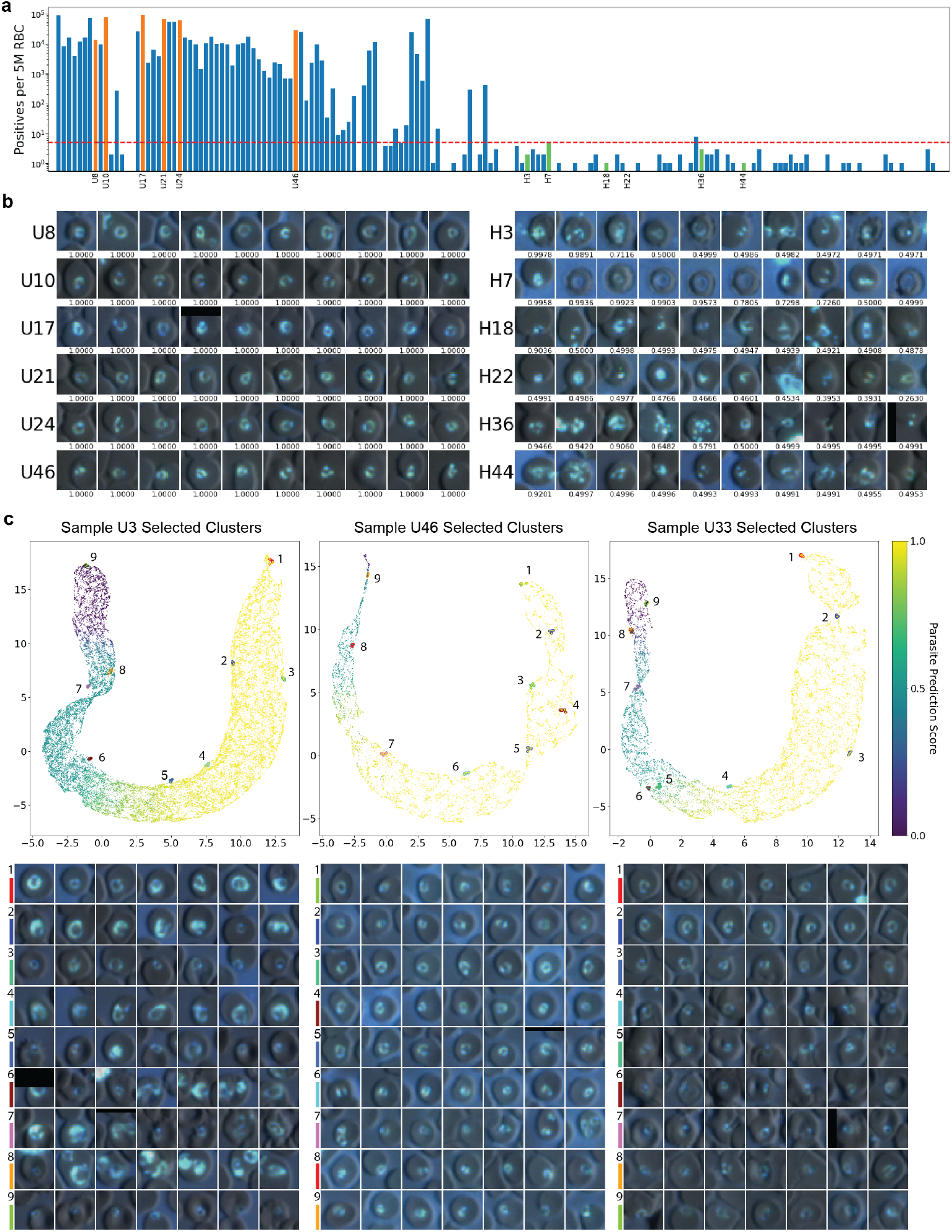
Quantification of parasitemia and morphological analysis of DAPI-stained malaria parasites and distractors in patient samples. **a**, Parasitemia quantification across samples from Uganda (starting with ‘U’) and from Stanford Blood Center (starting with ‘H’) using preliminary ResNet-34 model, assuming threshold of parasite prediction score = 0.667. **b**, Representative spots images (10 highest parasite prediction scores) from malaria positive (orange in **a**) and malaria negative (green in **a**) samples, with corresponding parasite prediction scores underneath images. **c**, Uniform manifold approximation and projection (UMAP) visualizations of manually annotated parasites from three patient samples, colored by parasite prediction scores. Representative fluorescent spot images from selected clusters in their respective UMAP plots demonstrate the diversity of detected parasites.

A major bottleneck in training our models was the need to annotate a large number of images. To address this challenge, we developed an interactive annotation tool. The workflow begins by having the user annotate a small subset of images, followed by training a preliminary model on these annotations. This trained model is then used to rank and sort the images, making subsequent annotations faster and more straightforward. The user can retrain and refine the model iteratively based on newly acquired annotations until the desired accuracy is achieved and/or number of images are annotated. (supplementary video 7). Using this tool we were able to annotate more than 34,000 images in less than 2.5 hours. To facilitate multi-user annotation by clinicians, we further developed a web-based annotation platform (Fig. S7). Finally, we created a web-based dashboard for interactive data visualization and exploration (Fig. S8).

### Machine learning model generalization through re-training

The initial classifier was trained on a dataset from 10 healthy donors and 10 malaria positive individuals (Fig. 4a). When tested on an unseen dataset comprising 164,509 spot images from 9 malaria positive slides (from 9 individuals) and 10,995,017 spot images from 129 negative slides (from 38 healthy donors), the model showed a suboptimal sensitivity of 80.3%, despite having a specificity of 99.993% (Fig. S2 a, c). Using this initial model, many parasites in the testing dataset were misclassified, as observed by the high false negative rate in the malaria positive slides (columns in Fig. 4c) to maintain 5 false positives per microliter in individual negative slides (rows in Fig. 4c). Examples of false positive and false negative images for some samples using this initial model are shown in Fig. 4d.

**Figure 4.**
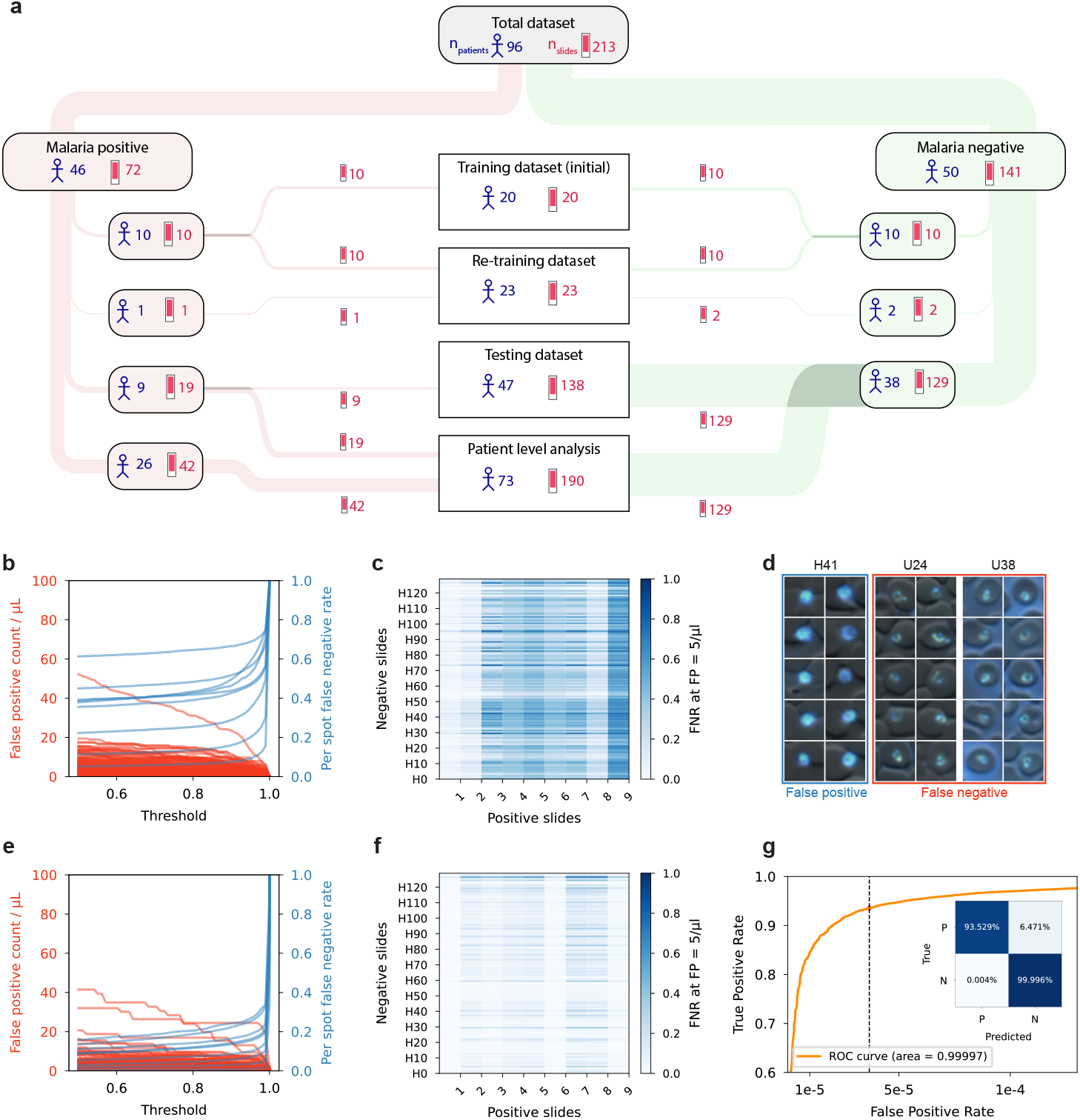
Machine learning classification of malaria parasites - slide and spot level. **a**, The distribution of patient/donor samples and slides into different datasets used for machine learning; *e*.*g*. the total dataset consists of 96 individuals (46 malaria positive patients, 50 malaria negative donors) and 213 slides (72 positives, 141 negatives). *n*_*patients*_ = number of patients or donors; *n*_*slides*_ = number of slides. **b**,**e**, Before and after re-training: false positive count per microliter and per spot false negative rate v.s. per spot parasite prediction score threshold. Each line represents one patient slide. **c**,**f**, Before and after re-training: per-spot false negative rate in malaria positive slides (columns) when the false positive rate in malaria negative slides (rows Hi) is equal to 5 parasites per microliter. **d**, Representatives false positive and false negative spots using the initial model. **g**, Receiver operating characteristic (ROC) curve for spot level detection with area under ROC curve 0.99997. Inset shows confusion matrix for parasite prediction score threshold of 0.5. ROC curve for initial model and full-range ROC curve for final model (g) shown in Fig. S2.

Recognizing that some variations of distractors and parasites may not be present in the limited original training set, we expanded the training set with 1,433,269 spot images from 2 healthy donors and 2,865 spot images from a malaria-positive individual (Fig. 4a). Re-training with this new set improved the true positive rate to 93.5% while maintaining average false positives per microliter below 5/*µ*L (Fig. 4e-g and Fig. S2). The marked improvement from re-training suggests that the model could achieve good performance in real-world settings through continuous and adaptive learning.

Moreover, when trained/tested on images using different channels, the model performs best when using all RGB channels (fluorescence) and DPC, confirming the utility of taking advantage of the spectral shift in DAPI staining for malaria parasite classification (Fig. S3).

### Patient-level analysis of machine learning performance

The performance of the machine learning algorithm at the patient level was tested on an independent dataset of 73 patients/donors and 190 slides (Fig. 4a), which had been excluded from the original training and re-training datasets. The predicted parasitemia for 61 slides from 35 malaria positive patients ranged from 16 parasites/µL to around 130,000 parasites/µL, whereas the predicted parasitemia for 129 slides from 38 malaria negative donors ranged from 0 to 41 parasites/µL (Fig. 5a,b). A patient-level threshold (corresponding to the cutoff for the number of suspected parasites per microliter for designating the case as malaria positive or negative) can be selected to balance the sensitivity and specificity trade-off for different clinical scenarios (Fig. 5c). In our current patient-level analysis dataset, the sensitivity and specificity for a threshold of 15 parasites/µL are 100% and 97.4%, respectively. Changing the threshold to 20 parasites/µL resulted in sensitivity and specificity of 97.1% and 100%. Since the sensitivity number is parasitemia dependent, we simulated the performance for 3 different parasitemia (Fig. 5h,i), assuming Poisson distribution and average object-level sensitivity of 0.92 and mean false positives of 3.47 per microliter (mean from 38 healthy donors). The overall diagnostic performance improves as the number of RBCs screened increases (Fig. S4), highlighting the utility of high-throughput screening for low-parasitemia cases. The estimated limit of detection (LoD) [63] for the ResNet-18 model is around 12 parasites per microliter (calculations in Methods section).

**Figure 5.**
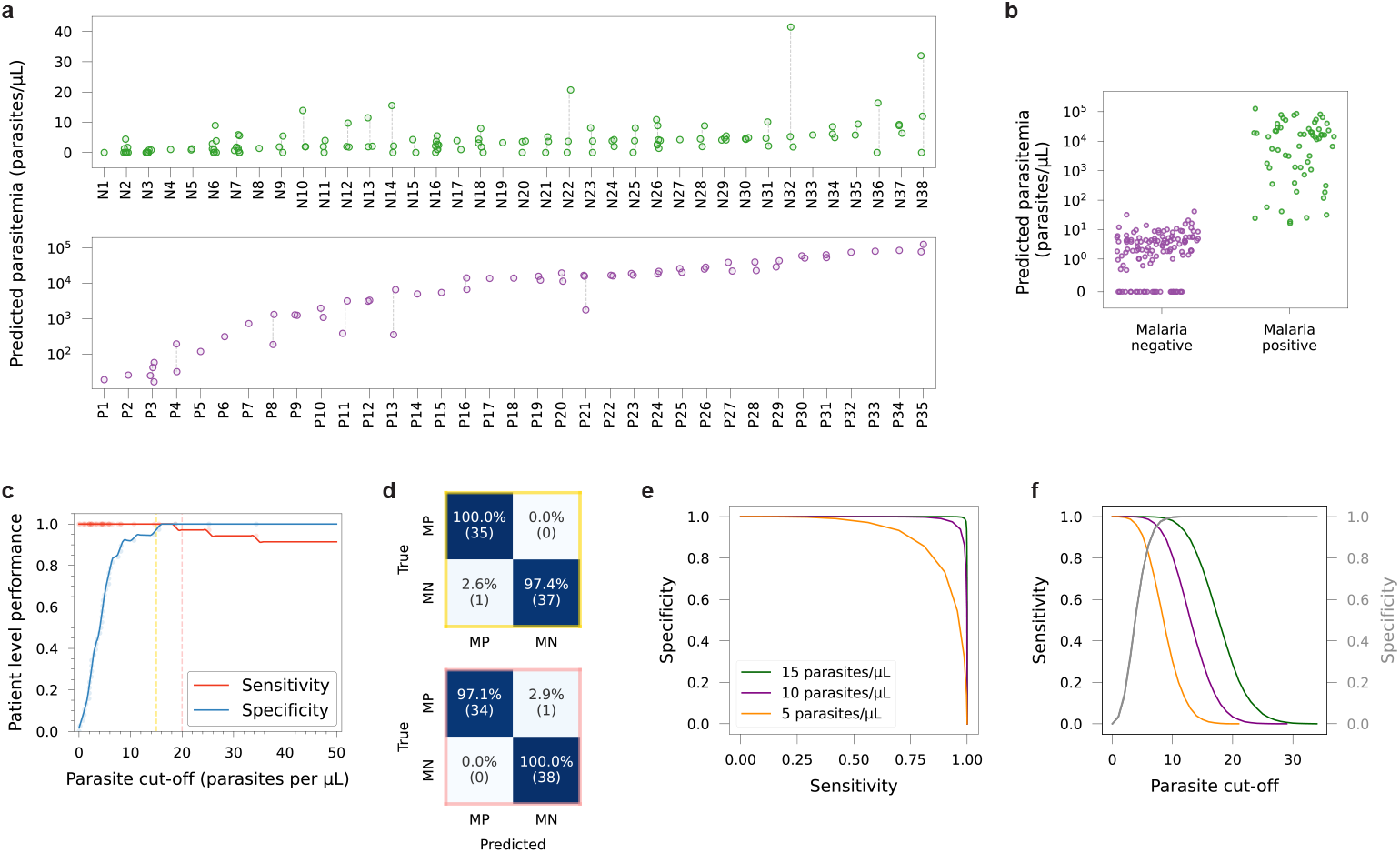
Detection of malaria at the patient level. **a**, Predicted parasitemia variation of each individual considered in the independent testing dataset for patient-level analysis, with unique data points corresponding to different slides; top plot shows data used for patient-level analysis for 38 malaria negative donors (129 slides), and bottom plot shows data for 35 malaria positive individuals (61 slides). N = negative; P = positive. **b**, Predicted parasitemia for all slides from malaria positive patients and malaria negative donors in the testing dataset for patient-level analysis. **c**, Patient-level performance (sensitivity and specificity) for 73 unique individuals (35 malaria positive, and 38 malaria negative); dashed vertical lines correspond to parasite cut-off values where confusion matrices were calculated. **d**, Confusion matrices for arbitrarily chosen parasite cut-off values of 15 parasites/*µ*L (yellow) and 20 parasites/*µ*L (pink). MP = malaria positive; MN = malaria negative. **e**,**f**, Simulated results of specificity and sensitivity for parasitemia of 5, 10, and 15 per *µ*L assuming 5 million red blood cells screened (plots for other RBC counts in Fig. S4).

## Discussion

Here we report the implementation of an AI-enabled, modular, and fully automated imaging platform that uses a simple glass slide interface and can be adapted for multi-disease diagnostics. Recognizing the importance of robustness and that the device eventually needs to be manufactured in certified facilities to meet standards and regulations, we freed ourselves from the constraints associated with DIY approaches and chose to use established components (*e*.*g*. cross-roller tables/stages and machine vision cameras) and fabrication method (CNC machining). The overall design, hardware modules made available from this implementation, and open-source software serve as the basis for rapidly developing cost-effective automated imagers configured and optimized for a wide range of diagnostic applications.

The imager presented in this work uses a 20x objective (can be easily switched to other magnifications with or without an objective turret), 405 nm for epi-illumination with a 435 nm long-pass emission filter and a 12MP color camera, making it directly suitable for many other diagnostic applications (e.g. cervical cancer screening, histopathology, detecting Mycobacterium tuberculosis in sputum samples with Auramine O or DMN-Tre [64]). The use of standard glass slides instead of proprietary cartridges and the associated low-cost, widely available consumables/reagents combined with the capabilities to image tens of thousands of slides per year could also help offset the upfront device cost in low-resource settings.

Applying the imaging platform to the diagnosis of malaria, we developed a solution that can determine parasitemia with a high degree of automation and high throughput (Fig. S5, Fig. S6, and supplementary videos 3 and 5). Before further technical development (e.g. cloud infrastructure for quality control) and regulatory clearance, human review of predicted parasite images and summary statistics should be part of the workflow. Ultimately, we envision a workflow requiring minimal user training, helping to alleviate the global shortage of skilled microscopists and enabling broader access to reliable malaria diagnostics [2, 10].

Furthermore, malaria control and elimination requires identification of human infectious reservoirs. While gametocyte carriage is subclinical, gametocytemic individuals comprise the parasite reservoir that leads to infection of mosquitoes and local transmission [65]. In one study, it has been shown that the transmission rate only increases significantly when the gametocyte density reaches 200 per *µ*L [66]. In another study, it was found that the most infectious individuals had an estimated density of 10 or more gametocytes per *µ*L, and that about 45–75% of all mosquito infections in their feeding assay are caused by individuals with total parasite densities below 100 parasites per *µ*L but gametocyte densities above 10 gametocytes per *µ*L [67]. In the same study, high-quality research-grade microscopy identified more than 90% of infectious *P. falciparum* carriers in high-transmission settings and two of three infectious carriers in a low-transmission setting. Due to the distinct morphological features, gametocytes can be unambiguously identified using our imaging platform at low magnification (Fig. S10). With scanning speed of 1.5 million red blood cells per minute (0.3 *µ*L blood per minute), detection limit below 1 gametocyte/*µ*L is likely achievable with scan time of less than 5 minutes. Since slide processing follows standard procedures already used in the field and reagent cost is less than $0.05 per test, with tools that help make consistent blood smears with minimal training required [61], our solution can be well suited for large scale surveys and epidemiology research studies.

While we use nucleic acid stains for sensitive detection of *P. falciparum* parasites in thin blood smears, different probes that are specific to a set of pathogens can also be utilized. The past decades have seen much development of pathogen-specific probes [68, 69, 64, 70]. Being low-cost and highly configurable, Octopi has the potential to help enable the widespread use of these new probes in low-resource settings.

There are several limitations in this pilot study. During both the training and testing phases, ambiguous samples from positive patients were excluded. In future studies fluorescent in situ hybridizations (FISH) can be used to provide ground truth labels. Although most of the negative slides from healthy donors were sourced from the Stanford Blood Center — raising the possibility of overfitting to this specific population — the model successfully classified unseen samples from healthy Ugandan donors, maintaining false positives below 5/µL. This suggests that overfitting is not a significant concern in the current dataset. Due to the availability of samples, currently the models are only trained for *P. falciparum*. Identification of other Plasmodium species will be developed as the next step. Our current approach also assumes a standard 5 million RBCs per µL, which may not account for variations in hematocrit due to factors such as sex, and age. Furthermore, the accuracy of parasitemia prediction was not assessed in this study due to the lack of a proper gold standard for parasitemia quantification, such as WHO expert level microscopy or qPCR. Studies are on-going to address most of the limitations by building larger clinical data sets with reference readouts.

## Materials and Methods

### Design of the microscope

Octopi 2.0 has evolved (Fig. S5) from proof-of-concept prototypes to custom built hardware for manufacturing. Latest Octopi 2.0 is designed to be easily configured for different applications due to its modular hardware, use of standard glass slides and an open-source Python-based software. The current configuration includes a lead screw stepper XY stage (60 mm × 60 mm travel with 2.54 mm pitch) and a lead screw linear actuator Z stage (7 mm travel with 0.3 mm pitch). A custom LED matrix array containing 128 programmable LEDs is used for transillumination, enabling bright-field microscopy, differential phase contrast imaging, and dark-field microscopy. The imaging optical train consists of an infinity corrected objective lens (*e*.*g*. Olympus 20×/0.5 for malaria), a machine vision lens (f = 50 mm for malaria) as tube lens and a CMOS camera (*e*.*g*. MER2-1220-32U3C, Daheng with Sony IMX226 sensor for malaria). For imaging DAPI-stained malaria parasites, epifluorescence excitation is provided by a 405 nm LED (M405L4, Thorlabs). A custom controller, including LED and stepper motor drivers, controls the illumination, motion and hardware trigger of the camera.

### Software with real-time processing

We developed a real-time malaria detection system that integrates hardware control, image processing, machine learning, and an interactive PyQt5-based user interface. For each run, the software scans a user-defined region on the blood smear. Prior to image acquisition, an autofocus calibration routine identifies the optimal focus range by scanning across a predefined z range and evaluating image sharpness. The optimal focus is determined for a few points in the scan region to generate a focus map for image acquisition. Then, a set of three images, including two bright-field images (left half and right half illuminated) images and one fluorescence image, are acquired for each location. The images are processed by deep learning models for cell segmentation and object classification. For each field of view, results including acquired bright-field and fluorescence images, processed differential phase contrast (DPC) images, segmentation masks, list of fluorescent spots and corresponding parasite scores (after classification), are stored for display and optionally uploaded to cloud storage. For efficient resource utilization and high-throughput performance, the software uses Python’s multiprocessing framework to coordinate and parallelize tasks, ensuring that processes such as image acquisition, DPC processing, segmentation, fluorescent spot detection, classification, data saving, and GUI presentation run concurrently. For each FOV with roughly 9,000 RBCs, image acquisition and processing takes around 1 second using a laptop with i7 CPU and RTX4070 mobile GPU.

### Image processing

For each FOV, a bright-field image pair, including images captured using illumination from the left-half and right-half of the programmable trans-illumination LED array (*I*_*L*_ and *I*_*R*_), is used to create a differential phase contrast (DPC) image *I*_*DPC*_ = 0.5 +(*I*_*L*_ − *I*_*R*_)*/*(*I*_*L*_ + *I*_*R*_). Cells in the DPC images are segmented using Cellpose (or modified M2U-Net for real-time processing), in order to count the number of red blood cells (RBCs) and to filter out fluorescent spots that are outside of RBCs.

The DAPI-stained fluorescent spots are detected using Laplacian of Gaussian (LoG) post background removal via white top-hat filtering with a structuring element of 17 × 17 pixels.

### Image annotation

For the initial training dataset, we manually annotated 55,755 positive spot images from 10 positive patients’ slides and randomly selected 744,187 negative spot images from 10 healthy donors’ slides. For the initial testing dataset, we manually annotated 161,643 positive spot images from 9 positive patients’ slides and selected 10,995,017 spot images from 129 slides of 38 healthy donors. During the re-training, we augment the original training dataset by additionally annotating 2,865 spots from one positive patient and 1,433,269 spots from 2 negative individuals. The training set and testing set are sourced from mutually exclusive patients. The patient-level data set is a superset of the testing set that includes the positive patient slides without annotations, consisting of 129 slides from 38 healthy donors from Uganda and Stanford Blood Center, and 61 slides from 35 malaria positive patients from Uganda.

### Deep learning based parasite detection

Following fluorescent spot detection and cell segmentation, spots outside red blood cells were omitted for classification. The filtered spots were then cropped to a standardized size of 31×31 pixels across four channels (one greyscale DPC channel and three fluorescence R,G,B channels). These cropped spot images served as input for neural network classifiers (ResNet [71]), which output a parasite prediction score, *i*.*e*. the softmax likelihood from the classifier’s final layer. The object-level detection of parasites occurs after the initial spot detection, thus red blood cells without fluorescent spots are not considered for classification. This addresses a common limitation mentioned in [72], and our approach significantly reduces the imbalance between distractors and positive objects compared to typical techniques used with Giemsa-stained slides.

For residual network structure [71], we adjusted the initial convolutional layer to have a kernel size of 3, a stride of 1, a padding of 1, and an input channel size of 4. For training, we used the Adam optimizer [73], with a batch size of 32, a learning rate of 0.001, cross-entropy loss, and training over 10 epochs. We selected ResNet-34 for initial model and ResNet-18 for re-training model. We utilized a validation set, which was 12.5% of the training set, to monitor performance and selected the model checkpoint with the lowest validation loss for final evaluations. We used PyTorch [74] version 2.1 to construct the model. All the training and testing were done on laptops with i7 CPU and RTX4070 mobile GPU.

### Patient-level analysis

Patient-level analysis was performed on a dataset containing 129 slides from 38 malaria negative donors from Uganda and Stanford Blood Center, and 61 slides from 35 malaria positive patients from Uganda. The dataset was independent from those used in training and retraining, *i*.*e*. data from the same patient or donor (including all slides per patient/donor) did not repeat between testing and training (or retraining) datasets. The ground truth diagnoses confirming malaria positive/negative in samples from Uganda was determined through microscopic examination. Samples from Stanford Blood Center were presumed to be malaria negative since malaria is not endemic in the region. The performance metrics (sensitivity, specificity and accuracy) were calculated at the patient-level for 73 patients/donors, by aggregating data from multiple slides for each individual. The cellular level or spot threshold for the machine learning (ResNet-18 model) was 0.5.

### Limit of detection calculation

The limit of detection (LoD) is defined as the parasitemia level at which the algorithm can consistently distinguish between positive and negative samples. To estimate the LoD for our malaria detection algorithm, we used the formula introduced in [63], equation (3). As defined in [63], we computed the following two metrics: 1) *Per-patient object sensitivity* ***S*** is the fraction of parasites of a positive sample that are correctly labeled for each patient (malaria positive); and 2) *Per-patient False Positive Rate* ***F*** is the number of distractors mislabeled as parasites per microliter for each healthy donor (malaria negative), namely FPs/µL. The LoD is estimated by the mean of per-patient object sensitivity *µ*(***S***) (0.924 for 9 positive patients), and the standard deviation of the per-patient FPR *σ*(***F***) (3.472 FPs/µL for 38 healthy donors), with the following formula:

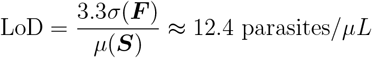

Thus, the estimated LoD is approximately 12 parasites*/µL*. Several assumptions were made in this calculation, as introduced in [63]. We assumed that the FPR follows a Gaussian distribution. The choice of the factor 3.3 is to enforce 95% specificity on negative samples as described in [75].

### Blood samples from healthy donors and from patients diagnosed with malaria

De-identified blood samples (whole blood) from healthy anonymous donors were obtained from the Stanford Blood Center in BD Vacutainer blood collection tubes with K2 ethylenediaminetetraacetic acid (EDTA) or heparin anticoagulant. De-identified methanol-fixed finger prick blood smears from patients diagnosed with/without malaria were provided by Infectious Disease Research Collaboration, Kampala, Uganda. The study protocols were approved by the Uganda National Council of Science and Technology (HS 2700), the Makerere University School of Medicine Research and Ethics Committee (2019-134), the University of California, San Francisco Committee on Human Research (19-28606), and the Institutional Review Boards at Stanford University (52977).

### Preparation and staining of blood smears

Smears of blood were fixed by dipping in absolute methanol for 30 seconds. Fixed smears were incubated with 5 *µ*g/mL DAPI solution for 2 minutes, washed in water, and air dried in the dark. DAPI solution was purchased from Biotium (catalog # 40043) and diluted to 5 *µ*g/mL. Stained blood smears were kept in dark before imaging.

### Data availability

The data for this study will be made publicly available before publication of the final version of the manuscript.

Code, software and models are publicly available. The software and firmware for Octopi is available in https://github.com/cephla-lab/squid, the code for the graphical user interface (GUI) with real-time processing and identification of malaria parasites is available in https://github.com/octopi-project/octopi-malaria-gui/. Processing code is available at https://github.com/octopi-project/octopi-malaria-processing.

## Acknowledgements

We thank Ellen Yeh and her lab including Marta Walczak and Katrina Hong for providing *P. falciparum* blood cultures. We thank Grant Dorsey from UCSF and Harriet Ochokoru, laboratory technician at Infectious Diseases Research Collaboration, Kampala, Uganda for providing de-identified patient blood smears via UCSF Malaria Eliminate program at the very beginning of the project. We thank Brieuc Cossic (Roche), Darvin Scott Smith (Stanford), Niaz Banaei (Stanford) for providing non-malaria samples. We thank Andres Garchitorena and Matt Bonds from PIVOT and Milijaona Randrianarivelojosia from Institute Pasteur, Madagascar and Sister Aquinas and staff of Swasthya Swaraj, Orissa, India and Vasundhara Rangaswamy (Stanford) for discussions on diagnostics needs in Madagascar and India. We thank David Giesbrecht, Jeff Bailey, Jenna Zuromski, Neeva Wernsman Young from Brown University, Jonathan J Juliano from UNC, Misago D. Seth and Deus Ishengoma from NIMR, Charles Delahunt from GH Labs for discussions on all aspects of the project. Finally, we thank all members of Prakash Lab for feedback and comments on the manuscript.

## Author contributions

H.L. and M.P. designed the instrument and overall research. H.L. built and characterized the instrument. H.L. collected data with assistance from M.P., L.V.F, J.P, K.M, B.O., H.SM. Heguang.L. and P.S.. H.L. developed models for spectral imaging. L.F.V, M.V. W.O. and K.M. developed the neural network for red blood cell segmentation. H.L., L.F.V. and M.V., R.B. and Heguang.L. developed processing pipelines and processed the data. M.P, P.S., J.P., E.L. and H.SM. performed field testing and usability studies. E.L. and H.L. developed new driver stack for the system. R.B. designed the new LED matrix for illumination. J.N., B.G., P.J and M.P provided oversight and feedback in design of field studies. H.L., Heguang.L., P.S and M.P. wrote the manuscript with feedback from all authors.

## Data and materials availability

All data necessary for interpreting the manuscript have been included. Additional information may be directly requested from the authors.

## List of Supplementary Materials

**Fig. S1**. Effect of DAPI stored in the dark at room temperature for 1 year.

**Fig. S2**. Receiver operating characteristic (ROC) curves and confusion matrices for initial ResNet-34 model and re-trained ResNet-18 model.

**Fig. S3**. Model performance using different channels (DPC only, greyscale+DPC, green+DPC, RGB+DPC).

**Fig. S4**. Simulation of sensitivity and specificity as a function of number of red blood cells.

**Fig. S5**. Evolution of Octopi over the years.

**Fig. S6**. Octopi scanning software user interface and program structure.

**Fig. S8**. Online interactive dashboard showing parasitemia count, UMAP visualizations, and sample images.

**Fig. S7**. Spot image annotation tool.

**Fig. S9**. Software architecture for real-time detection of malaria parasites.

**Fig. S10**. Examples of gametocytes imaged using Octopi.

**Movie S1**. Low magnification (top; reversed lens configuration from Octopi 1.0), and high magnification (bottom; Nikon Ti2 using a 20x/0.75 objective with 395/25 nm excitation and 460/50 nm emission) image pairs from parasite clusters, showing fluorescence (left) and brightfield/DPC (right) images. Link to video: https://youtu.be/Tj-CzrIxG1s

**Movie S2**. Low magnification (top; reversed lens configuration from Octopi 1.0), and high magnification (bottom; Nikon Ti2 using a 20x/0.75 objective with 395/25 nm excitation and 460/50 nm emission) image pair from platelet clusters, showing fluorescence (left) and brightfield/DPC (right) images. Link to video: https://youtu.be/hVb869jSlvo

**Movie S3**. Operation of the Octopi 2.0 platform and demonstration of real-time detection of malaria *P. falciparum* parasites. Link to video: https://youtu.be/u6E4ttEXp4E **Movie S4**. Differential phase contrast (left), fluorescence (middle) and overlay of DPC and fluorescence images of parasites, captured by Octopi. Link to video: https://youtu.be/s9KFhXwMv0c

**Movie S5**. A deep zoom image (overlay of fluorescence and differential phase contrast) of a patient slide scanned on the latest system. The image can be viewed online at https://bit.ly/3DWDhlw. Link to video: https://youtu.be/MMIUU75BFcM

**Movie S6**. Sample images of different clusters from the visualization of the uniform manifold approximation and projection (UMAP) of 10 patients. Exploration of different parasite morphologies is enabled by interactively selecting different clusters or regions in the UMAP. Link to video: https://youtu.be/Sf7YgDTTcbk

**Movie S7**. Example video of interactive annotation tool to annotate multiple spots and active learning tool for retraining. Link to video: https://youtu.be/OOX5M05uiLk

