## Supplementary File for "Octopi 2.0: Open and Scalable Microscopy Platform for AI-powered Diagnostic Applications"

March 21, 2025

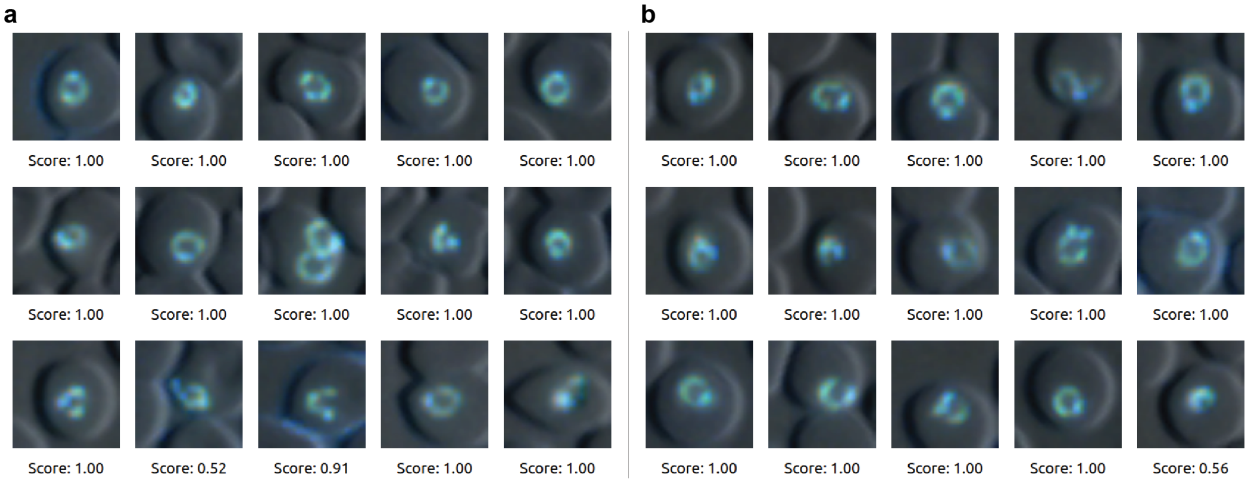


**Figure S1 Effect of DAPI stored in the dark at room temperature for 1 year.** **a**, Example images of DAPI-stained parasites for sample stained using fresh DAPI solution **b**, Example images of DAPI-stained parasites for sample (from the same patient) stained using DAPI solution stored in the dark at room temperature for 1 year, showing negligible effect on staining quality. Numbers below the images are parasite prediction scores provided by the model.

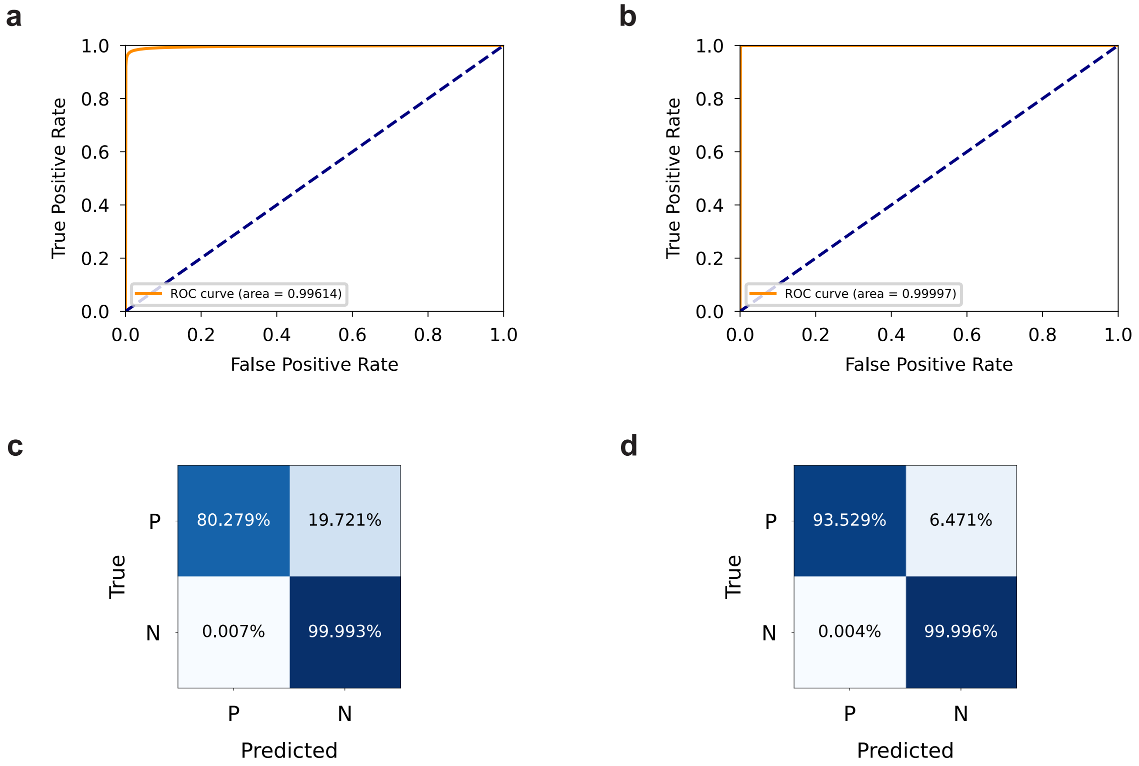


**Figure S2 Receiver operating characteristic (ROC) curves and confusion matrices for initial ResNet-34 model and re-trained ResNet-18 model.** **a**, ROC curve for initial ResNet-34 model, corresponding to Fig. 4 b,c. **b**, ROC curve for ResNet-18 model after re-training, corresponding to Fig. 4 e-g. **c**, Confusion matrix at a threshold of 0.5 for the initial ResNet-34 model. **d**, Confusion matrix at a threshold of 0.5 for the ResNet-18 model after retraining. Fig. 4 g shows a subset of ROC curve in b and shows the confusion matrix in d.

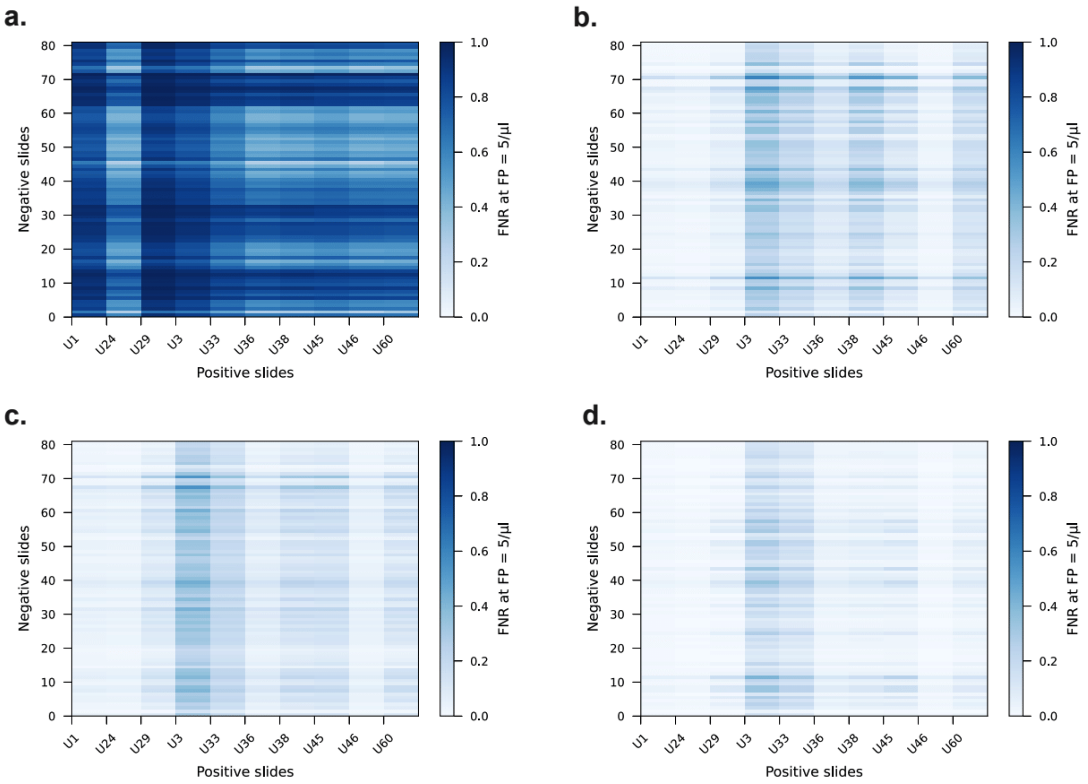


**Figure S3** **Model performance using different channels.** Model training with the same datasets but different channels shows the performance of models trained with channel-rearranged images. Heatmaps show the resulting FNR for positive datasets with threshold levels determined by negative datasets to achieve FPs count of 5 per microliter. Darker shades represent higher FNR. **a**, Model trained with images with differential phase-contrast (DPC) channel only. **b**, Model trained with grayscale channel (Pixel value = 0.299 * R + 0.587 * G + 0.114 * B) and DPC channel only. **c**, Model trained with green fluorescence channel and DPC channel only. **d**, Model trained with all channels (RGB+DPC).

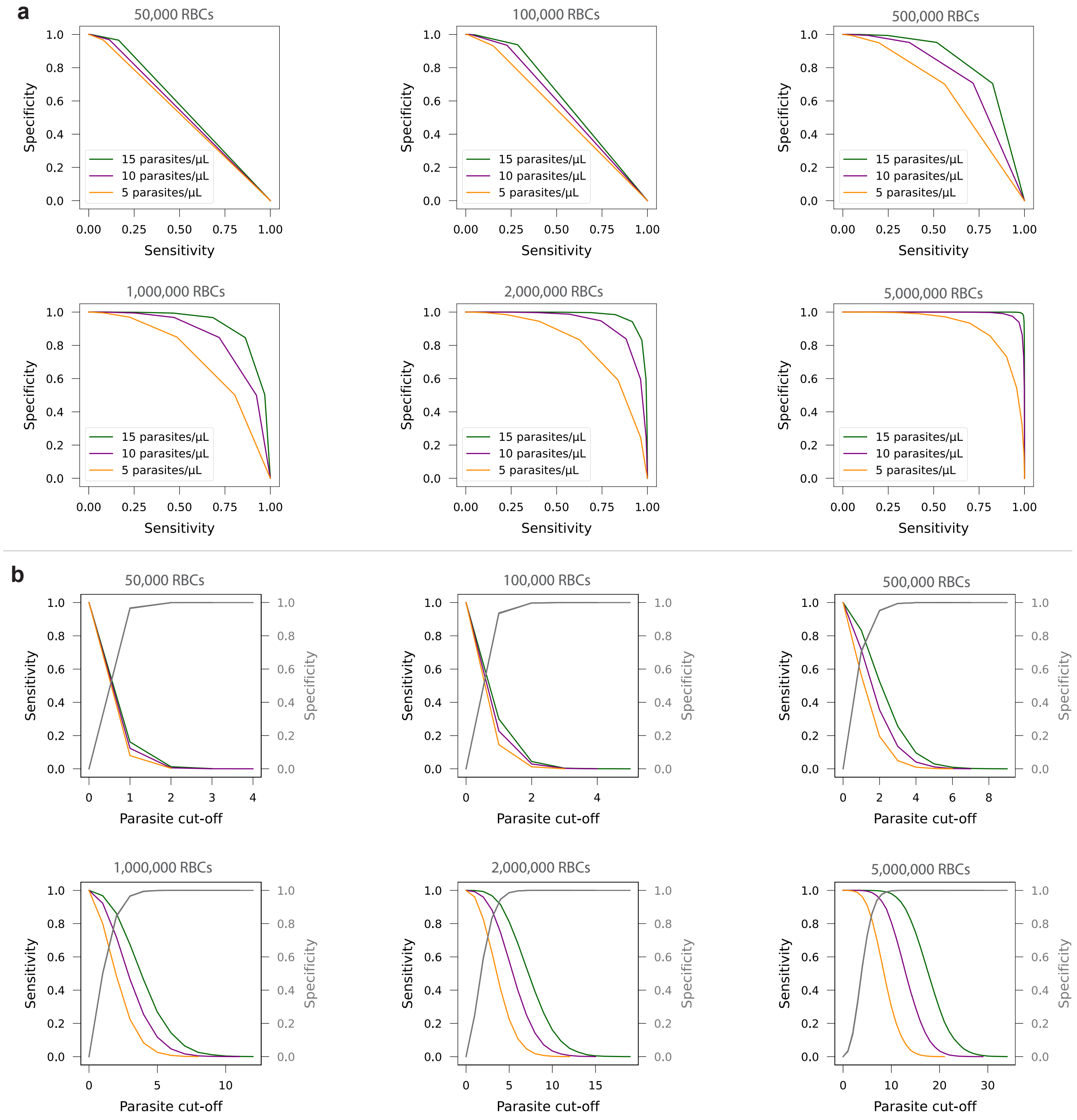


**Figure S4 Simulation of sensitivity and specificity as a function of number of red blood cells.** **a**, Receiver operating characteristic (ROC) curves for different parasitemia (5, 10 and 15 parasites/$\mu L$ - indicated by orange, purple and green curves, respectively) and different numbers of RBCs assumed in the simulation (50k, 100k, 500k, 1M, 2M, and 5M), showing that the overall diagnostic performance increases as the number of red blood cells screened increases. **b**, Sensitivity and specificity as a function of parasite cut-off for different numbers of RBCs.

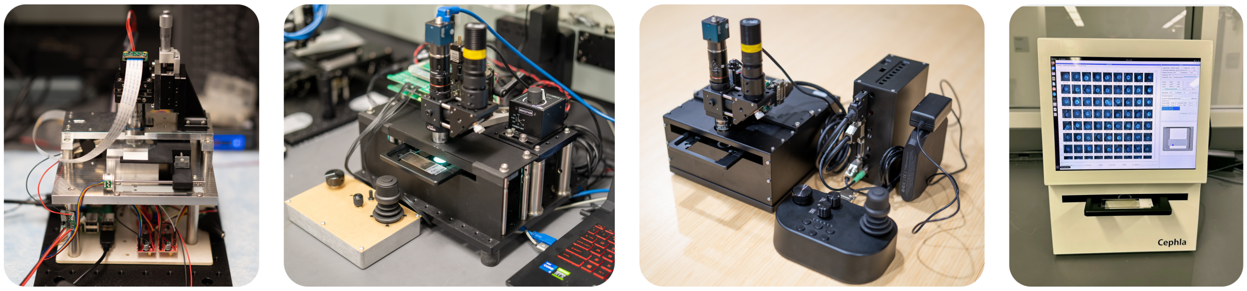


**Figure S5** **Evolution of Octopi.** Different versions of Octopi over the past few years, showing development of hardware and the concept of an integrated diagnostic device.

a

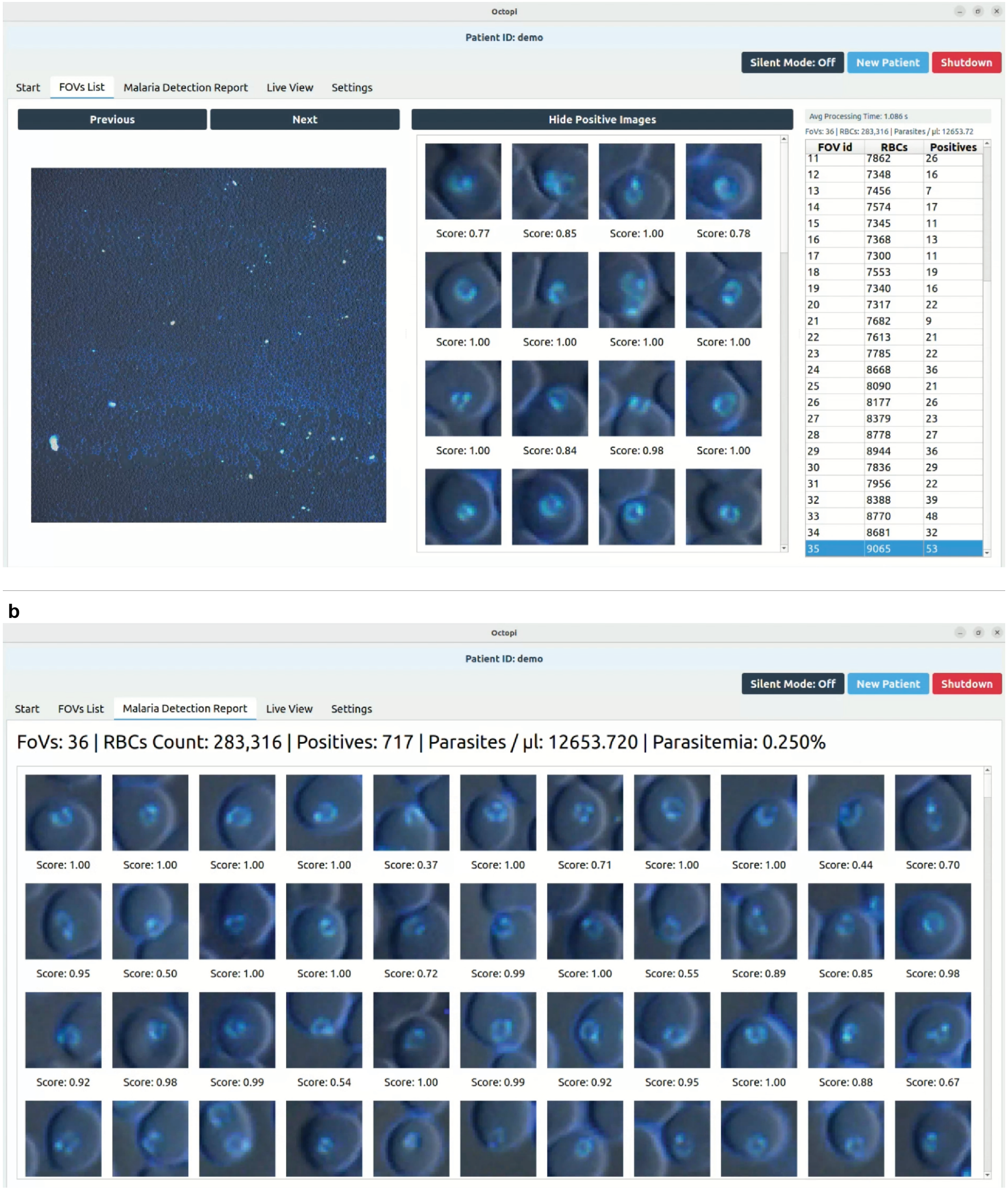


**Figure S6 Octopi scanning software user interface and program structure.** **a**, Software window showing a field of view, predicted parasites or positive images, and a list of FOVs with RBC count and number of predicted positive cells. **b**, Summary or detection report of all the scanned FOVs.

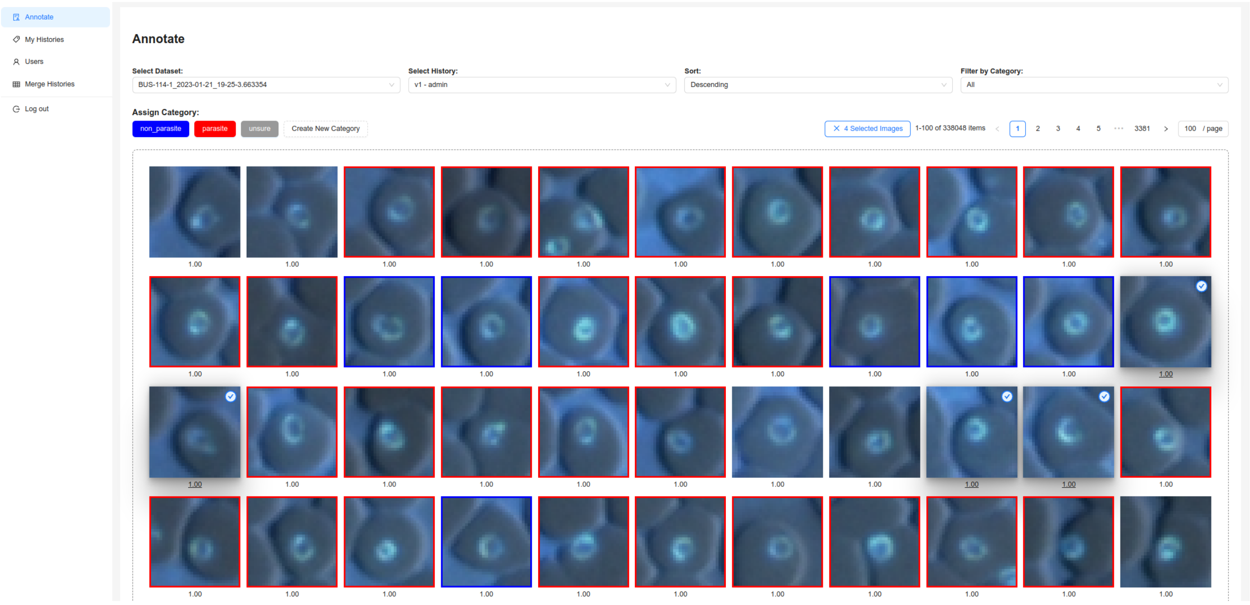


**Figure S7 Spot image annotation tool.** Online interactive tool for annotating detected fluorescent spot images into different categories: parasites, non-parasites, unsure, or new category. Note: annotations arbitrarily selected for demonstration purposes only

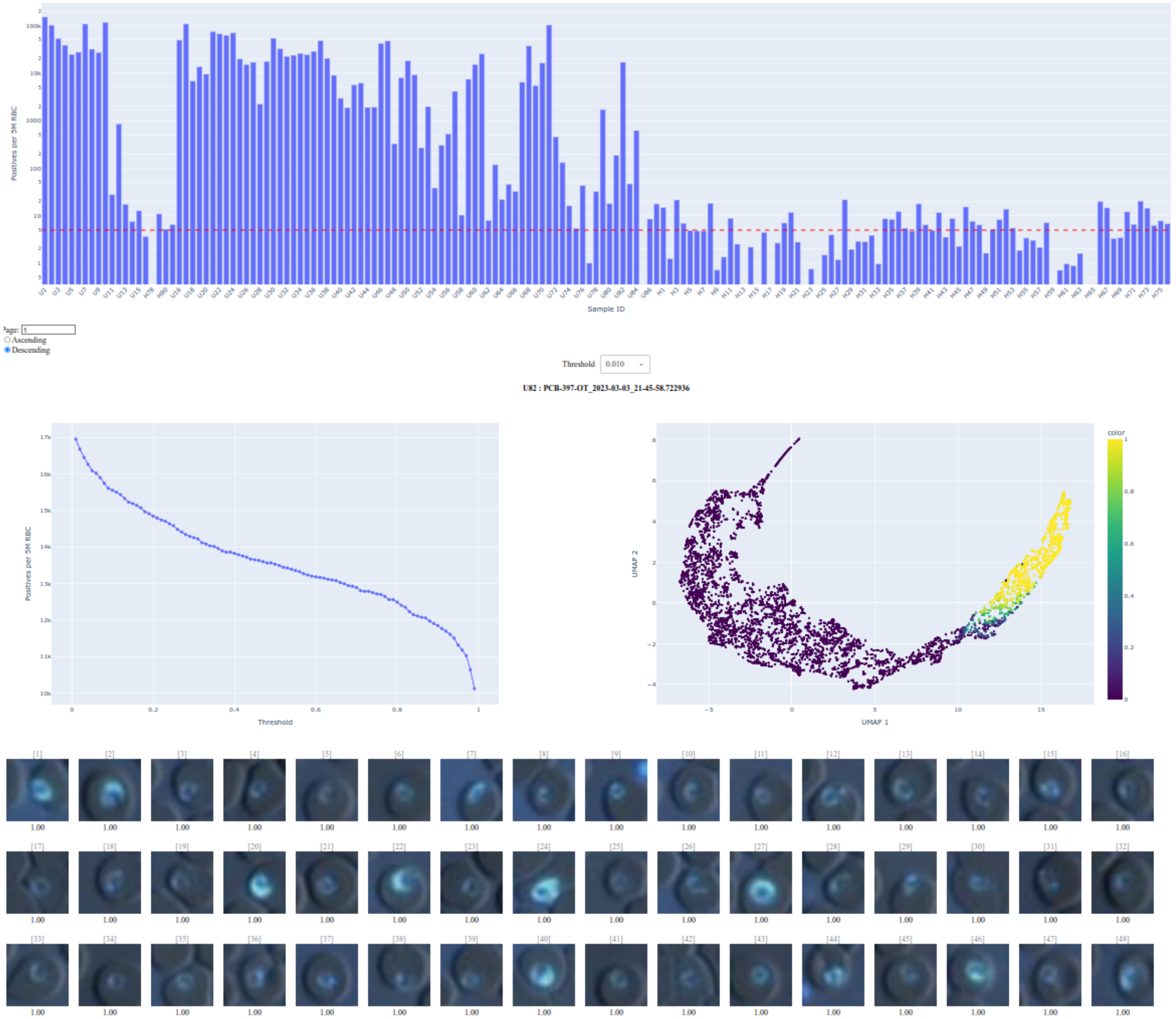


**Figure S8** **Online interactive dashboard.** A dashboard to visualize the parasitemia count, UMAP visualization of the image features, and sample images with parasite prediction scores. The threshold can be adjusted to visualize its effect on the number of predicted parasites or the predicted parasitemia

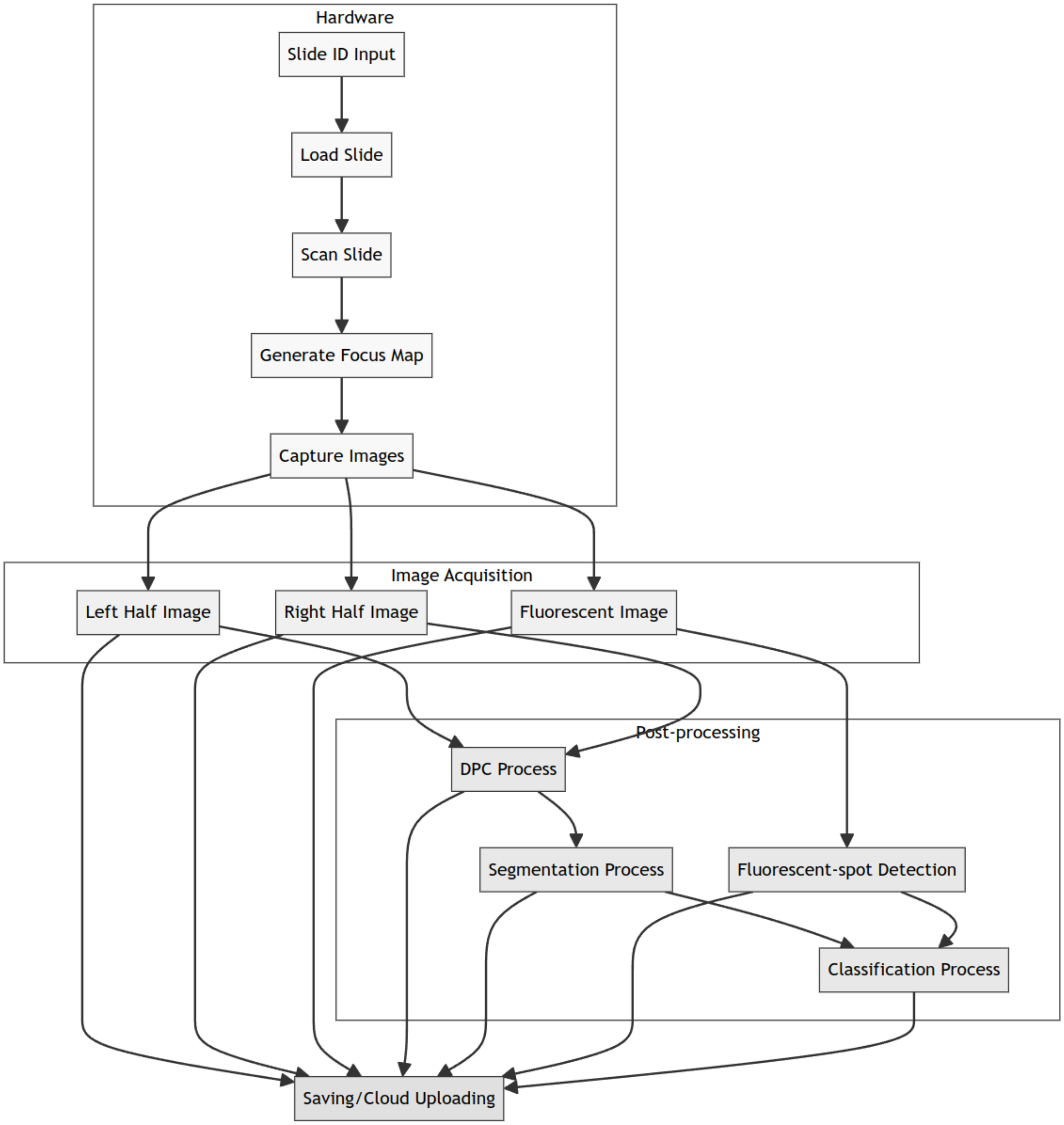


**Figure S9 Software architecture for real-time detection of malaria parasites.** Hardware interfacing, image acquisition and image-processing pipelines for malaria detection

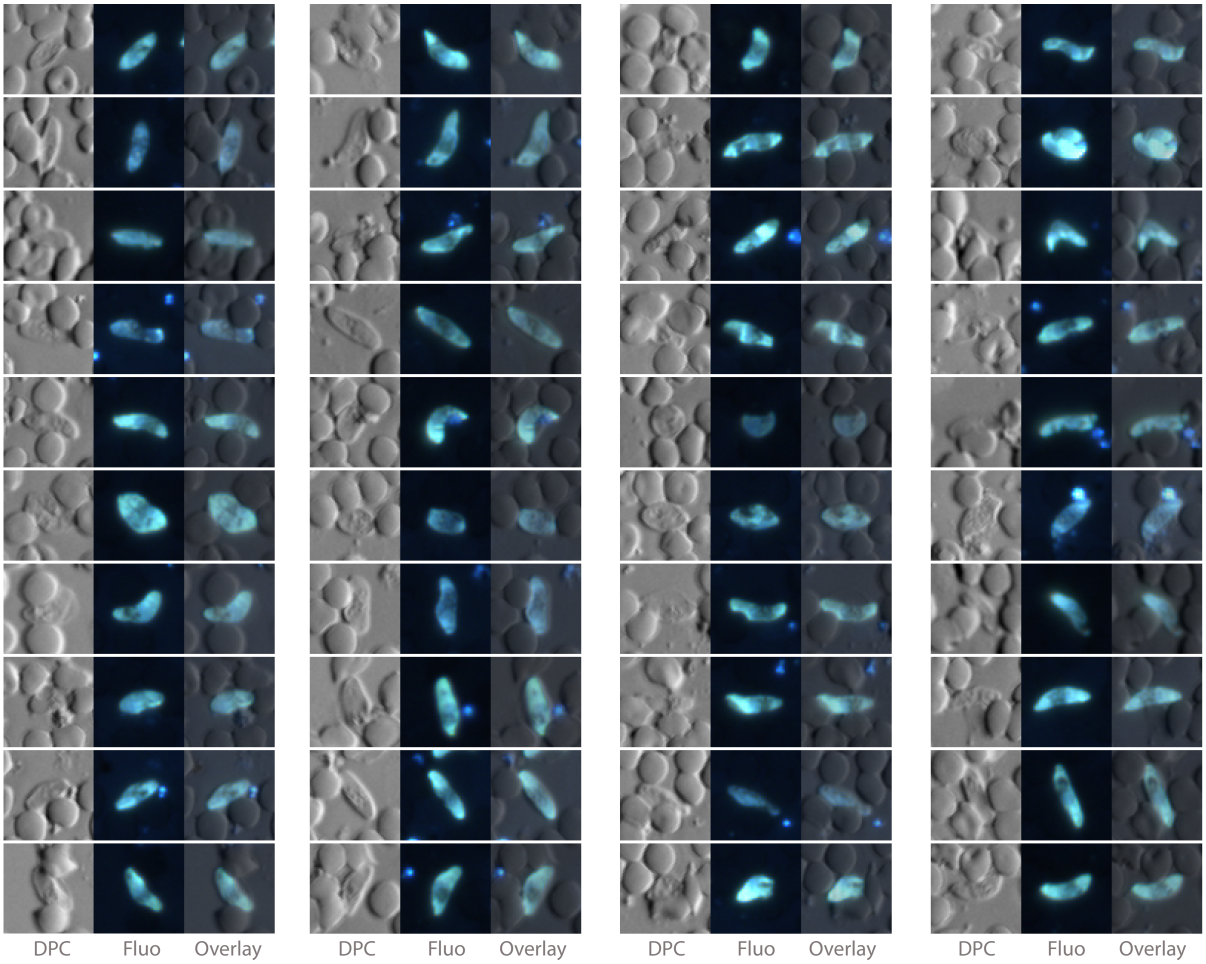


**Figure S10** **Examples of gametocytes imaged using Octopi.** Differential phase contrast (DPC), fluorescence (Fluo) and overlay of DPC and fluorescence (Overlay) images of DAPI-stained cultured gametocytes imaged using Octopi.

### Supplementary videos

**Movie S1.** Low magnification (top; reversed lens configuration from Octopi 1.0), and high magnification (bottom; Nikon Ti2 using a 20x/0.75 objective with 395/25 nm excitation and 460/50 nm emission) image pairs from parasite clusters, showing fluorescence (left) and brightfield/DPC (right) images. Link to video: https://youtu.be/Tj-CzrIxG1s

**Movie S2.** Low magnification (top; reversed lens configuration from Octopi 1.0), and high magnification (bottom; Nikon Ti2 using a 20x/0.75 objective with 395/25 nm excitation and 460/50 nm emission) image pair from platelet clusters, showing fluorescence (left) and brightfield/DPC (right) images. Link to video: https://youtu.be/hVb869jSlvo

**Movie S3.** Operation of the Octopi 2.0 platform and demonstration of real-time detection of malaria *P. falciparum* parasites. Link to video: https://youtu.be/u6E4ttEXp4E

**Movie S4.** Differential phase contrast (left), fluorescence (middle) and overlay of DPC and fluorescence images of parasites, captured by Octopi. Link to video: https://youtu.be/s9KFhXwMv0c

**Movie S5.** A deep zoom image (overlay of fluorescence and differential phase contrast) of a patient slide scanned on the latest system. The image can be viewed online at https://bit.ly/3DWDhlw. Link to video: https://youtu.be/MMIUU75BFcM

**Movie S6.** Sample images of different clusters from the visualization of the uniform manifold approximation and projection (UMAP) of 10 patients. Exploration of different parasite morphologies is enabled by interactively selecting different clusters or regions in the UMAP. Link to video: https://youtu.be/Sf7YgDTTcbk

**Movie S7.** Example video of interactive annotation tool to annotate multiple spots and active learning tool for retraining. Link to video: https://youtu.be/00X5M05uiLk
